# Outbreak of COVID-19 among vaccinated and unvaccinated homeless shelter residents — Sonoma County, California, July 2021

**DOI:** 10.1101/2021.12.07.21267204

**Authors:** Aleksandr Bukatko, Mark N. Lobato, Emily Mosites, Cameron Stainken, Katheryn Reihl, Mojgan Deldari, John M. Bell, Mary Kate Morris, Debra A. Wadford, Kathleen Harriman, Sundari Mase

## Abstract

In July 2021, the Sonoma County Health Department was alerted to three cases of COVID-19 among residents of a homeless shelter in Santa Rosa, California. Among 153 shelter residents, 83 (54%) were fully vaccinated; 71 (86%) vaccinated residents had received the Janssen COVID-19 vaccine and 12 (14%) received an mRNA (Pfizer BioNTech or Moderna) COVID-19 vaccine. Within 1 month, 116 shelter residents (76%) received positive SARS-CoV-2 test results, including 66 fully vaccinated residents and 50 not fully vaccinated. 9 fully vaccinated and 1 unvaccinated were hospitalized for COVID-19. All hospitalized cases had at least one underlying medical condition. Two deaths occurred, one in a vaccinated resident and one in a non-vaccinated resident. Specimens from 52 residents underwent whole genome sequencing; all were identified as SARS-CoV-2, Delta Variant AY.13 lineage. Additional mitigation measures are needed in medically vulnerable congregate setting where limited resources make individual quarantine and isolation not feasible.

## Introduction

Outbreaks have occurred in homeless shelters throughout the COVID-19 pandemic (1). Furthermore, people experiencing homelessness have been reported to have a higher risk of hospitalization compared to the general population (2). As a result, layered prevention measures have been recommended in homeless service sites (3).

Here, we report an investigation of an outbreak of COVID-19 that occurred in a homeless shelter in Sonoma County, California in July 2021, among residents who were fully vaccinated as well as those who were not fully vaccinated.

## Methods

On Epi-week 27, 2021, three cases of COVID-19 were identified by local hospitals among residents of a 213-bed homeless shelter in Santa Rosa, California. At the time, 153 persons were residing in the shelter, where they were housed in six congregate dormitories. No new residents entered the shelter over the course of July. Among shelter residents, 83 (54%) were fully vaccinated. After an outbreak of COVID-19 in the same shelter in January 2021, the facility had spaced beds 6 feet apart and implemented a universal mask policy for staff members and residents. Routine testing was not part of the prevention measures already in place. Upon identification of the new cases, 73 shelter residents sleeping in the two dorms of the three index patients were immediately quarantined as a cohort because rooms for individual quarantine were not available. After further positive results were identified, all dorms were quarantined as cohorts. Quarantine adherence was reported to be variable. Residents and staff members were provided with KN95 and surgical masks, although staff reported inconsistent mask use among residents. Contact tracing was conducted to identify contacts within the shelter and for other locations visited. Exposed staff members who were not fully vaccinated were sent home to quarantine.

On epidemiology week 28, 2021, Sonoma County Health Department initiated symptom checks and twice-weekly SARS-CoV-2 nucleic acid amplification tests (NAAT; reverse transcription–polymerase chain reaction) for residents and staff members in collaboration with a partner hospital. Cases were defined as receipt of a positive SARS-CoV-2 NAAT result. Fully vaccinated was defined as persons who were ≥2 weeks after receiving the second dose in a 2-dose series (Pfizer-BioNTech or Moderna), or ≥2 weeks after receiving a single-dose vaccine (Johnson & Johnson [J&J]/Janssen). Vaccine breakthrough infections were defined as positive NAAT test results among persons who were fully vaccinated at the time of specimen collection. Four partially vaccinated residents (those who either did not complete the 2-dose series (for those receiving an mRNA vaccine) or for whom less than 2 weeks had elapsed since receipt of the final dose) were considered not fully vaccinated in the analysis. Reinfection was defined as a positive test result during this outbreak in a person who had a recorded positive test result >90 days earlier. Contacts were defined as any person within 6 feet of a COVID-19 patient for a cumulative total of ≥15 minutes over a 24-hour period, irrespective of mask use. Sonoma County Health Department staff interviewed residents with positive SARS-CoV-2 results to collect demographic and clinical information, including underlying medical conditions. Underlying conditions included those defined by CDC to be associated with risk for severe COVID-19 (https://www.cdc.gov/coronavirus/2019-ncov/need-extra-precautions/people-with-medical-conditions.html). Medical records for hospitalized persons were reviewed. Vaccination status was confirmed through California Immunization Registry records. Comparisons between residents who were vaccinated and those who were not fully vaccinated were performed using Fisher’s exact test due to small sample size.

Specimens from 52 residents underwent whole genome sequencing at the California Department of Public Health Viral and Rickettsial Disease Laboratory and Sonoma County Public Health Laboratory. Sequencing was done with Clear-Labs automation platform using Oxford-nanopore technology for sequencing and Guppy algorithm for basecalling. The consensus genomes generated from whole genome sequences (n=51 sequenced in 2 groups) underwent phylogenic analysis using the Ultrafast Sample placement on Existing tRees (UShER) bioinformatics application (4). NC_045512.2 (SARS-Ncov-2 isolate Wuhan-Hu-1) was used as the reference genome sequence for phylogenic analysis. For the purpose of easier differentiation between sublineages of the basal strain (the most common ancestor between the detected sequences), sequences in the study were given alphanumeric designations corresponding to further divergence from the basal strain (Appendix Figure 1, Appendix Table 1a). Serum samples from eight of 10 hospitalized persons were tested for total anti-SARS-CoV-2 nucleocapsid (N) antibodies immunoglobulin (Ig)M, IgG, and IgA (BioRad Platelia, California, USA), and by SARS-CoV-2 enzyme-linked immunosorbent assay to detect anti-SARS-CoV-2 IgG to viral spike (S), matrix (M), and N proteins, and for neutralizing antibodies (United Biomedical, Inc, New York, USA).

## Results

In a month between Epi-weeks 27 and 32, 116 (76%) residents received positive SARS-CoV-2 test results (Figure 1). The mean age of residents with positive test results was 52 years (standard deviation: 13 years), and 73 (63%) were male (Table 1). Among those for whom information on ethnicity was available (n = 142), 118 (83%) were non-Hispanic and 24 (17%) were Hispanic. Seventy-four residents (64%) had documented symptomatic disease. Data on existing medical conditions were available for 93 (80%) infected residents; 82 (88%) of whom had at least one condition associated with risk for severe COVID-19. Ten residents were hospitalized for COVID-19, including four who were admitted to an intensive care unit; two residents died. All 10 residents hospitalized for COVID-19 were ≥55 years old and had at least one underlying medical condition (seven had more than two conditions) including chronic obstructive pulmonary disease, diabetes, cardiovascular disease, and substance use disorders. Cancers and rare conditions were also noted.

**Figure 1.**
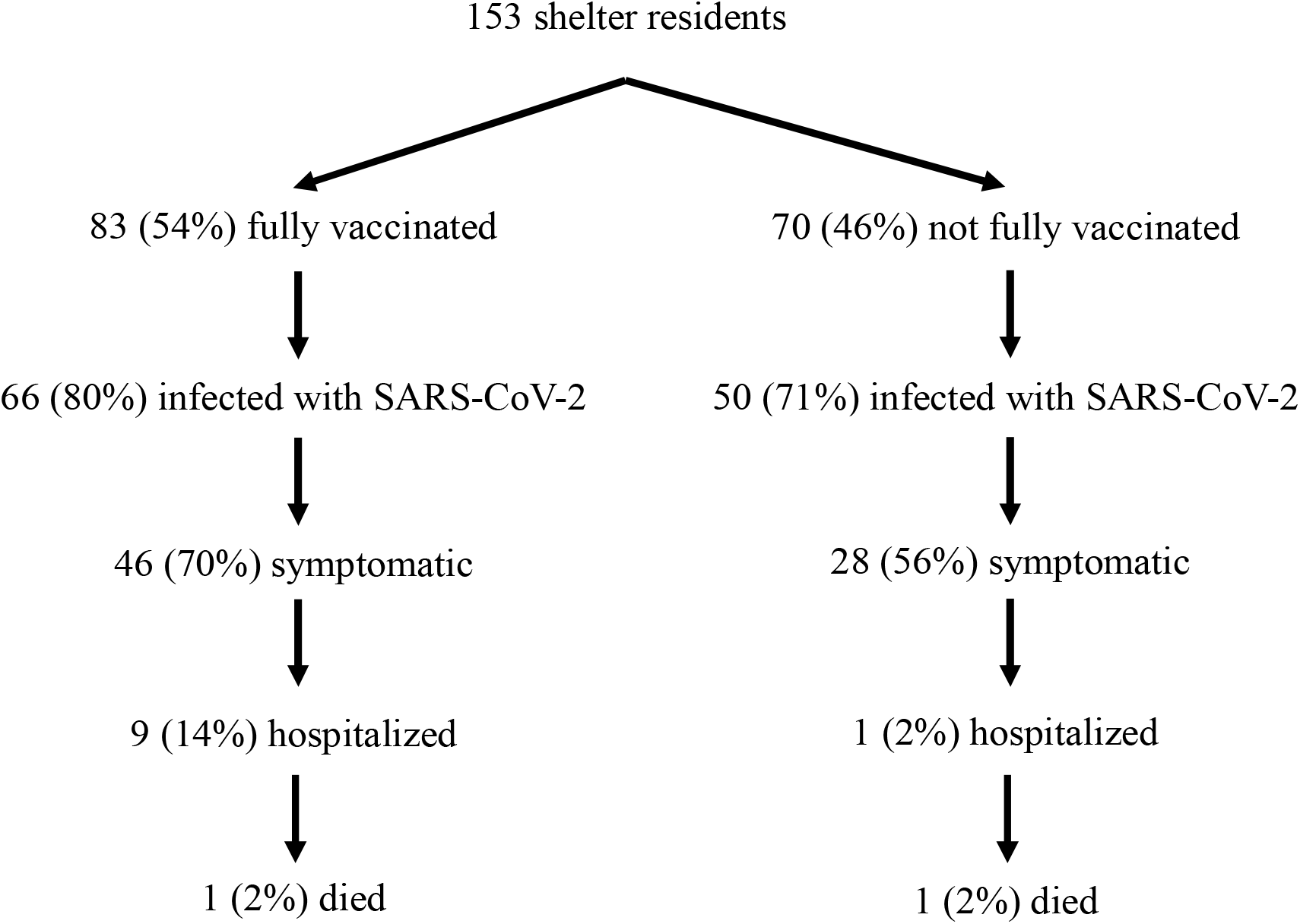
Vaccination status, infection, hospitalization, and death among residents of a homeless shelter during a COVID-19 outbreak — Sonoma County, California, July 2021. * Fully vaccinated included persons who were ≥2 weeks after receiving the second dose in a 2-dose series (Pfizer-BioNTech or Moderna), or ≥2 weeks after receiving a single-dose vaccine (Johnson & Johnson [J&J]/Janssen) ^†^Among those who received a positive test result for SARS-CoV-2 ^§^ Including 4 residents who were partially vaccinated

**Table 1.**
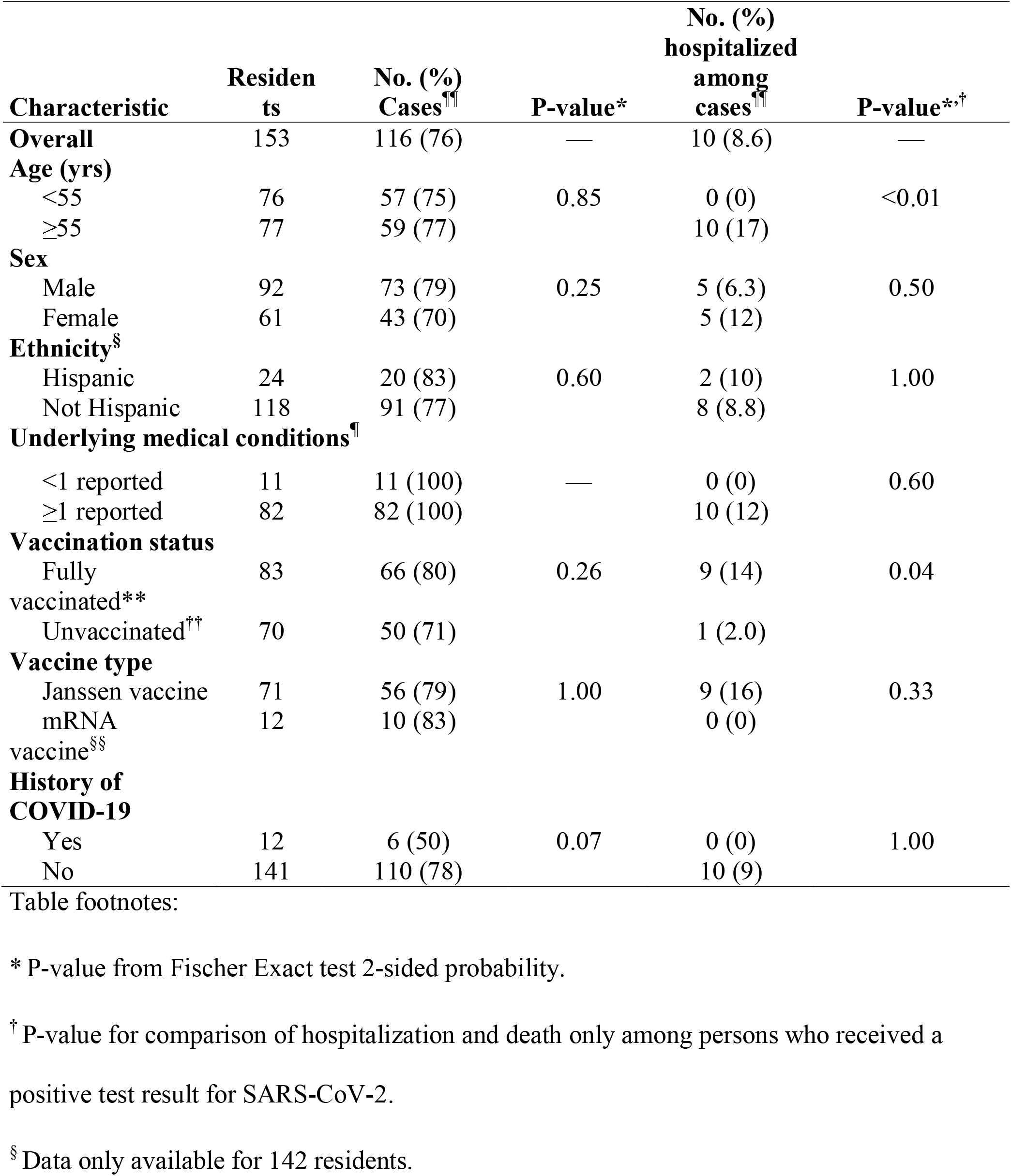
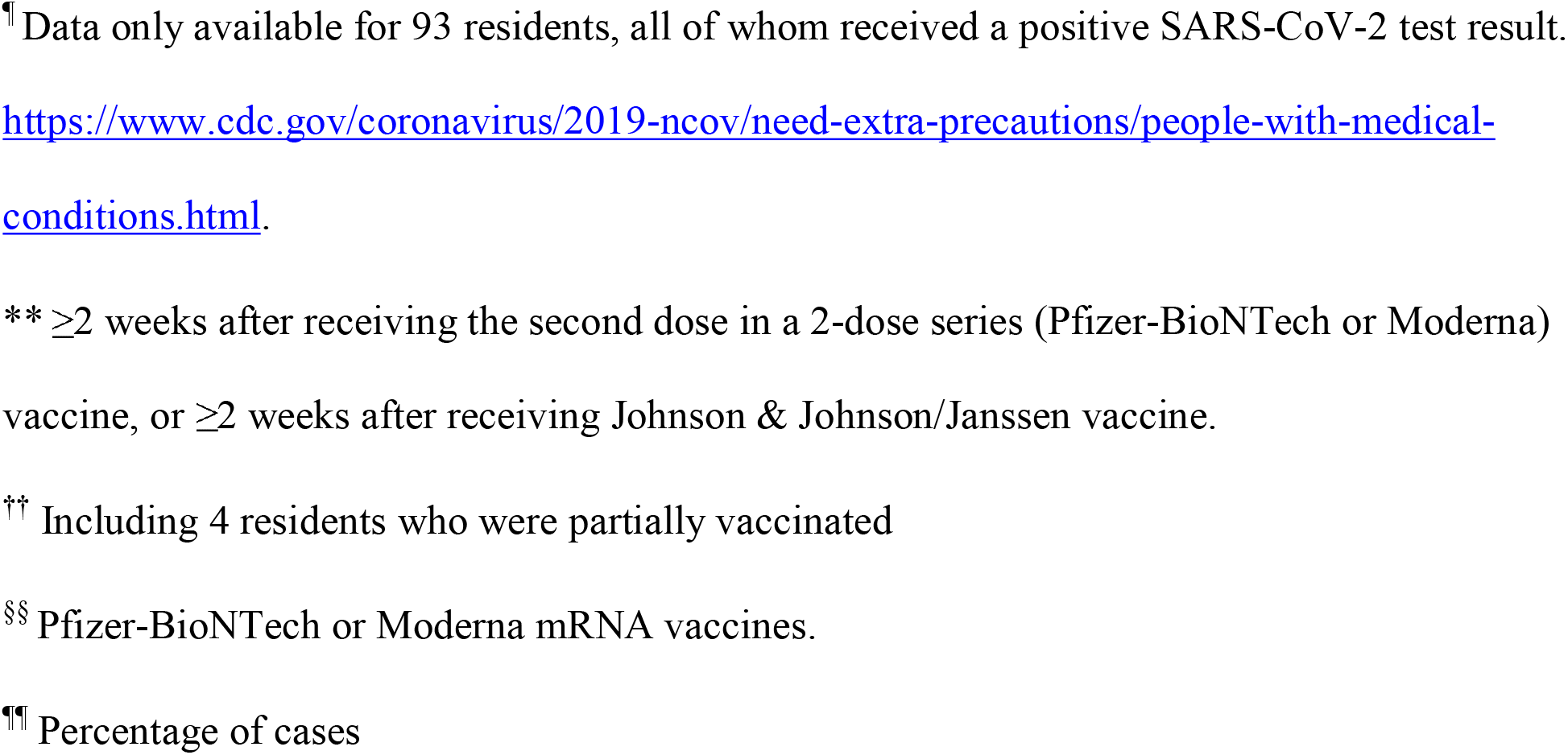
SARS-CoV-2 infections, hospitalizations, and deaths among residents of a homeless shelter — Sonoma County, California, July 2021.

All sequenced specimens were identified as SARS-CoV-2 (the virus that causes COVID-19) AY.13 lineage, a sublineage of the B.1.617.2 (Delta) variant. Fifty WGS samples from shelter residents were available for single nucleotide polymorphisms (SNP)-level phylogeny analysis and 14 substrains of AY.13 were detected (Appendix Table 1b). One substrain came from sample collected before the first systematic round of testing, 7 additional substrains were detected by first round of within-facility testing (with one dorm containing 6 and one dorm containing 4 different substrains). SNP-level sequencing analysis identified 6 top/bottom bunks where WGS results are available for both residents: 4/6 bunks contained discordant sequences, 2/6 bunks contained concordant sequences.

All eight available serum samples for hospitalized residents were antibody positive; however, one sample tested negative for IgG(S,N,M) and positive for total antibody(N); another sample was positive for IgG(S,N,M) and negative for total antibody(N).

Contact tracing at the shelter revealed additional epidemiologically linked NAAT– confirmed cases, including 2 cases among health care providers who worked at the shelter, 10 cases at the primary workplace of the health care providers (five staff members, one visitor, and four staff family members), one case in a delivery person, and eight cases in three encampments where shelter residents spent time.

Among the 116 shelter residents with positive SARS-CoV-2 test results, 66 (57%) were fully vaccinated; 56 (85%) of those received the Janssen vaccine. The mean interval since achieving fully vaccinated status was 10 weeks, including among those with positive test results. Five reinfections occurred during the outbreak among 12 persons with recorded past infection; three of the five reinfections occurred in persons who were fully vaccinated with Janssen COVID-19 vaccine. No patients with reinfection were hospitalized or died.

Janssen vaccine administered to shelter residents had 7 different lot numbers; 54% of vaccinated persons received one specific lot number. Positive test result was not associated with any lot numbers. The cohort was vaccinated by 19 different providers. Fifty-one percent were vaccinated by one particular provider, which was not associated with positive result compared to other providers. Among those vaccinated, 36% were vaccinated during a single event on March 20, 2021. However, that event was not associated with positive result compared to other administration events.

Residents aged ≥55 years were more likely to be vaccinated (47/77; 61%) than were those aged <55 years (36/76; 47%), but the difference was not statistically significant (p = 0.11). Residents with at least one underlying condition (55/82; 67%) were more likely to be vaccinated than were those without an underlying condition (3/11; 27%) (p = 0.02).

Infection occurred in 66 of 83 (80%) fully vaccinated residents and 50 of 70 (71%) residents who were not fully vaccinated (Table 1). Among the 74 residents with documented symptomatic disease, 46 (62%) were fully vaccinated. All 10 hospitalizations (nine in fully vaccinated residents) and both deaths occurred among residents aged ≥55 years. Three of four residents admitted to the ICU were fully vaccinated. Hospitalization was significantly associated with being fully vaccinated (p = 0.04). This association was not statistically significant after stratifying for age or having ≥2 underlying conditions. The type of vaccine received was not statistically significantly associated with infection or hospitalization. Neither stratifying by prior infection nor excluding persons who were partially vaccinated from the analysis meaningfully changed the results. Vaccine lot number, storage, and administering agency were unrelated to COVID-19 outcomes.**

## Discussion

This outbreak of SARS-CoV-2 AY.13, a sublineage of the B.1.617.2 (Delta) variant occurred in a homeless shelter and resulted in severe outcomes among residents who were not fully vaccinated as well as those who were fully vaccinated. In this setting, residents who were fully vaccinated became infected, were hospitalized, and died from COVID-19 during the outbreak. However, results should be interpreted with caution because of the unique characteristics of this setting.

This outbreak demonstrated a high attack rate (76%) which could be a result of quarantine being conducted as a cohort, increased transmissibility of the variant, and medical vulnerability of the resident population. The high diversity of subvariants at onset of outbreak suggests multiple introductions to the facility. Notable within-dorm diversity at first round of testing and discordance of viral genome sequences between a number of top/bottom bunk sample pairs suggests that interaction outside of sleep-time may have played a significant role in the infection spread.

Recent reports suggest waning immunity after vaccination *(5)* and lower efficacy of currently approved vaccines against infection with the Delta variant *(6,7)*. However, mRNA vaccine effectiveness against hospitalization has remained high and vaccinated persons have accounted for the minority of hospitalizations *(7,8)*. This outbreak in a unique setting with high risk of transmission showed results that differ from broader experience among vaccinated persons. This reinforces the importance of layered preventions strategies, including vaccination and non-pharmaceutical strategies (e.g. masking, physical distancing) to prevent SARS-CoV-2 transmission in settings at high risk for transmission such as homeless shelters. *(3)*

The reason that persons who were vaccinated had severe outcomes after infection during this outbreak is not clear. The serology results for eight of 10 hospitalized patients were positive for SARS-CoV-2, suggesting that these patients did mount an immune response. However, many shelter residents had medical conditions associated with more severe outcomes after infection with SARS-CoV-2, which might have placed those who were fully vaccinated at higher risk. Although we had limited ability to stratify data due to the size of the outbreak, confounding by age and underlying conditions likely led to the higher proportions of fully vaccinated persons who were hospitalized compared to those who were not fully vaccinated. Studies have also produced evidence that 1 dose and 2 dose regiments of currently available vaccines may yield insufficient protection in older age persons with no previous infection; additional vaccine doses may be beneficial for such persons to achieve adequate immune response to currently circulating variants *(9,10)*.

The findings in this report are subject to at least four limitations. First the sample size was small, limiting the power to determine the statistical significance of some differences of interest. Second, almost all residents received the Janssen vaccine, so it was not possible to determine whether vaccine type contributed to the findings here. Third, vaccine status was confirmed through the California Immunization Registry; persons who might have been immunized but not included in the registry could have been misclassified in this analysis. Finally, data on underlying conditions were not available for uninfected residents.

In this population at higher risk for severe COVID-19 living in a homeless shelter, SARS-CoV-2 infection, symptomatic disease, hospitalization, and death occurred among residents who were fully vaccinated and those who were not fully vaccinated. These findings reinforce the importance of layered prevention strategies, including appropriate isolation and quarantine, screening testing, physical distancing, and masking in addition to providing vaccination, prevent COVID-19 outbreaks in homeless shelters and similar congregate settings.

## Data Availability

All data produced in the present work are contained in the manuscript

---

**Appendix Figure 1.**
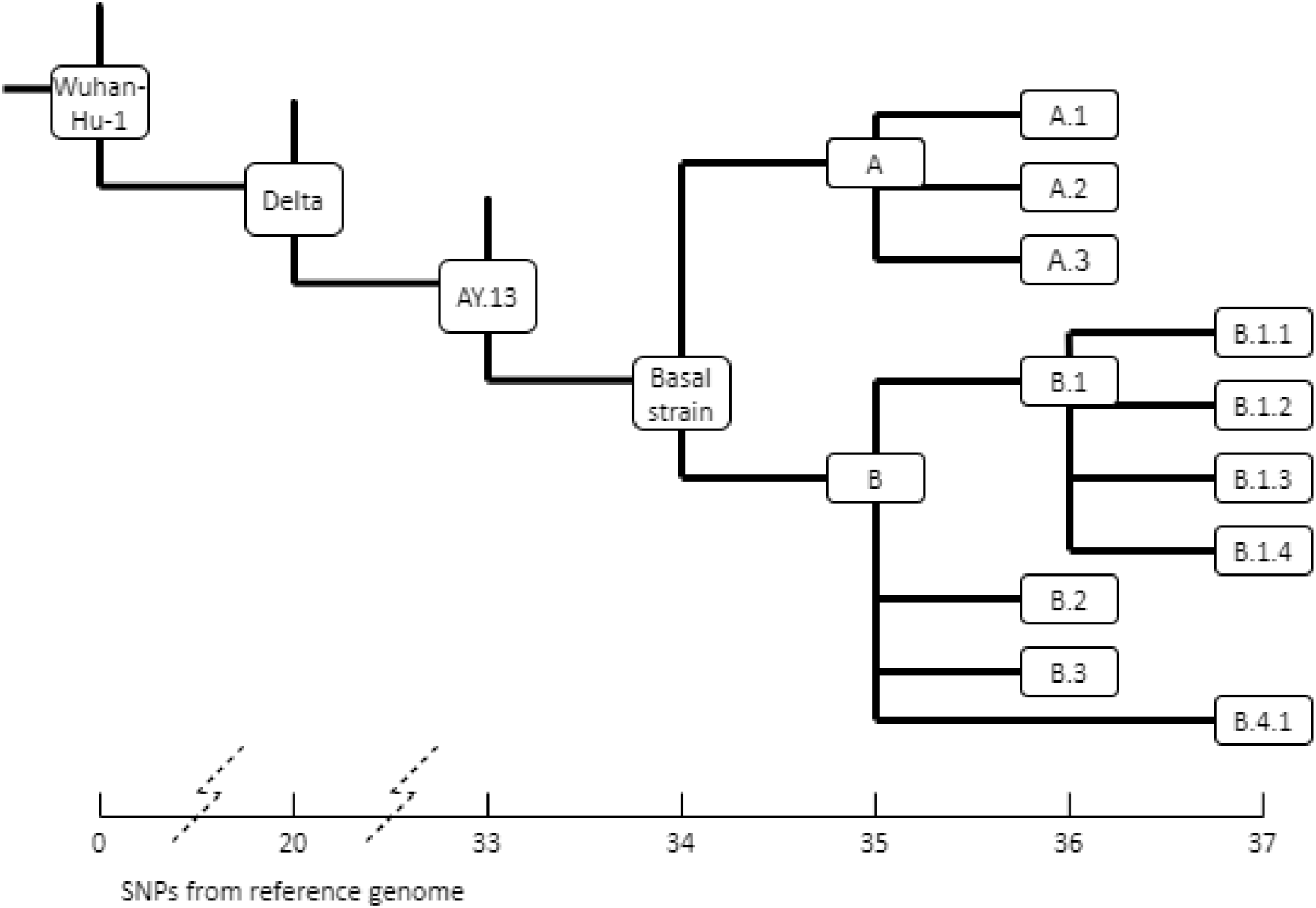
Phylogenetic tree using the Ultrafast Sample placement on Existing tRees (UShER) bioinformatics application. SNP (single nucleotide polymorphism). Basal strain for this outbreak was a 1 SNP divergence from AY.13 strain and a 34 SNP divergence from the reference genome of NCBI Sequence NC_045512.2 (SARS-Ncov-2 isolate Wuhan-Hu-1). Consensus genomes produced by analysis of sequencing results generated a phylogenetic tree using the Ultrafast Sample placement on Existing tRees (UShER) bioinformatics application.

**Appendix Table 1a.**
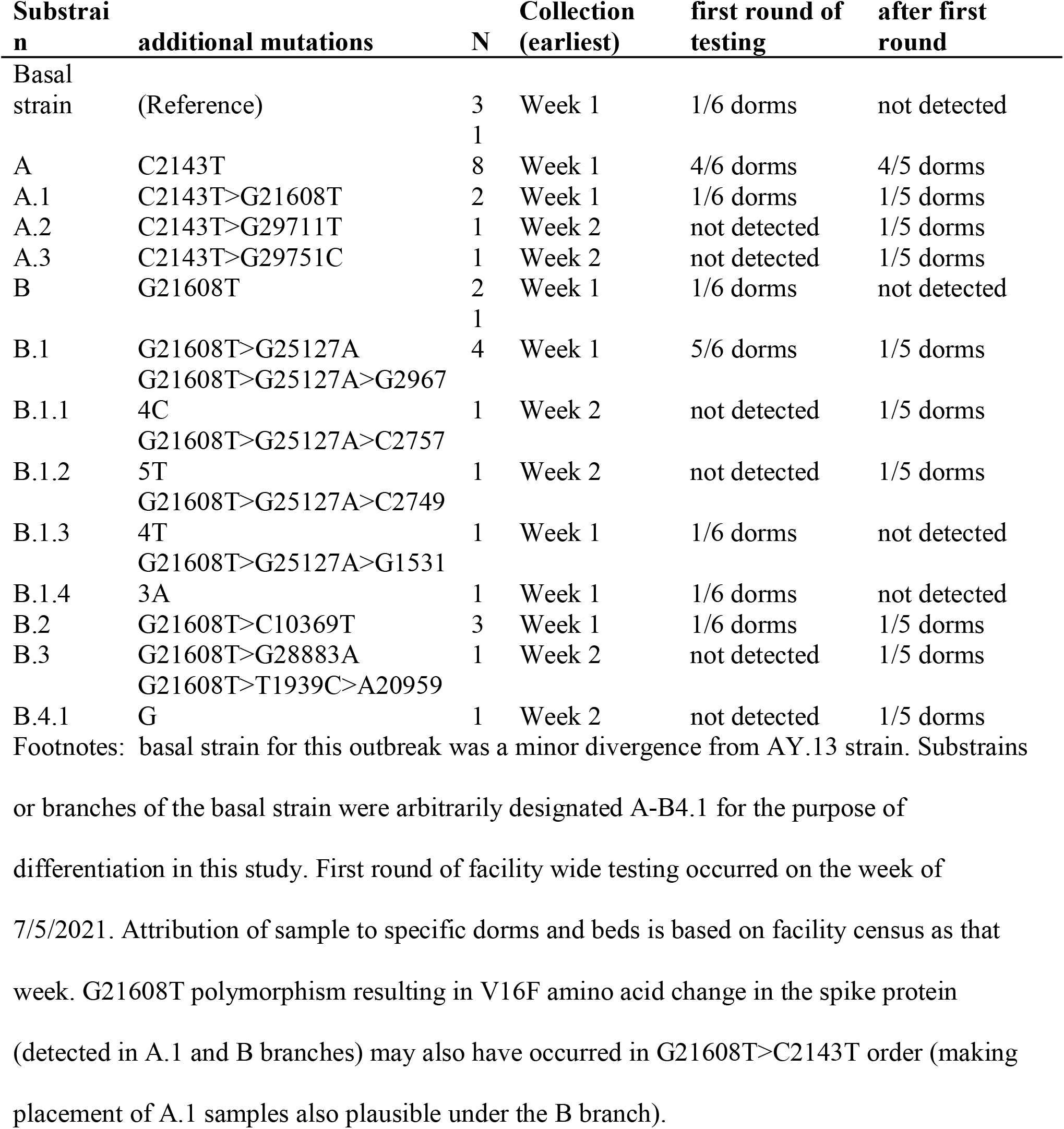
Phylogeny summary statistics.

**Appendix Table 1b.** Single nucleotide polymorphisms (SNP)

| **SNP** | **Amino Acid** |
| --- | --- |
| T1939C | none |
| C2143T | none |
| C10369T | none |
| G15313A | ORF1ab:A5017T |
| A20959G | ORF1ab:N6899D |
| G21608T | S:V16F |
| G25127A | S:V1189I |
| C27494T | ORF7a:P34L |
| C27575T | ORF7a:T61I |
| G28883A | N:G204R |
| G29674C | ORF10:X39Y |
| G29711T | none |
| G29751C | none |

## Acknowledgments

Jennielynn Holmes, staff, Sam Jones Hall; Jennifer Eid-Ammons, St. Joe’s Testing Team; Alan Powell, Daisy Cabrera, Kaiya Kramer, Maya Missakian, Lucinda Gartner, Rachel Rees, Outreach Team, Sonoma County Public Health Division. Maria Salas, Ruth Lopez, Leo Oceguera, Sutana Bethancourt, Sharon Messenger, Carl Hanson, CDPH/Viral and Rickettsial Disease Laboratory; Russell Corbett-Detig, University of California, Santa Cruz; Ashley Meehan, COVID-19 Homelessness Unit, CDC COVID-19 Response.

## Disclaimers

### Author Bio

Mr. Bukatko is an Epidemiologist at Sonoma County Department of Health Services, Santa Rosa, CA. His primary scientific interest is in molecular biology and immune response.

### Footnotes

This activity was reviewed by CDC and was conducted consistent with applicable federal law and CDC policy.^*^

## Footnotes

* See e.g., 45 C.F.R. part 46, 21 C.F.R.part 56; 42 U.S.C. §241(d); 5 U.S.C. §552a; 44 U.S.C. §3501 et seq.

